# The enactment of compassionate leadership by nursing and midwifery managers: Results from an international online survey

**DOI:** 10.1101/2020.09.16.20195115

**Authors:** Irena Papadopoulos, Steve Wright, Runa Lazzarino, Christina Koulouglioti, Magdeline Aagard, Özlem Akman, Lise-Merete Alpers, Paraskevi Apostolara, B. Julieta Araneda, Sylvia Biglete-Pangilinan, Orit Eldar Regev, Maria Teresa González-Gil, Christiana Kouta, Radka Krepinska, Małgorzata Lesińska-Sawicka, Miroslava Liskova, Lucero Lopez-Diaz, Maria Malliarou, Ángel Martín-García, S. Mara Muñoz, Małgorzata Nagórska, Roinah Nkhensani Ngunyulu, Sara Nissim, Line Nortvedt, Florinda Oconer-Rubiano, Cristina Oter-Quintana, Candan öztürk, Katalin Papp, Blanca Piratoba-Hernandez, Elena Rousou, Maria Ymelda Tolentino-Diaz, Valerie Tothova, Akile Zorba

## Abstract

**Aim:** To explore the views of an international sample of nursing and midwifery managers concerning attributes that they associate with compassionate management.

**Method:** A cross-sectional online survey. Using a snowballing sampling method, 1,217 responses were collected from nursing and midwifery managers in 17 countries. A total of complete 933 responses to a question related to which actions and behaviours indicated that a manager was exercising compassionate leadership were analysed for this paper. Firstly, content analysis of the responses was conducted, and secondly a relative distribution of the identified themes for the overall sample and for each participating country was calculated.

**Results:** Six main themes were identified describing the attributes of a compassionate leader: 1) *Virtuous Support*, 2) *Communication, 3) Personal Virtues of the Manager*, 4) *Participatory Communication*, 5) *Growth/Flourishing/ Nurturing*, and 6) *Team Cohesion*. The first three themes mentioned above collectively accounted for 63% of the responses, and can therefore be considered to be the most important characteristics of compassionate management behaviour.

**Conclusion:** The key indicators of compassionate management in nursing and midwifery which were identified emphasise approachability, active and sensitive listening, sympathetic responses to staff members’ difficulties (especially concerning child- and other caring responsibilities), active support of and advocacy for the staff team, and active problem solving and conflict resolution. While there were differences between the countries’ views on compassionate healthcare management, some themes were widely represented among different countries’ responses, which suggest key indicators of compassionate management that apply across cultures.

## INTRODUCTION

In the healthcare context, compassion has been defined as responding with kindness and sensitivity to the vulnerability and suffering of patients and their relatives - an altruistic concern for suffering coupled with the desire or motivation to alleviate it [1]. While incorporating empathy, there is also a behavioural, helping component in this definition - the desire or motivation to alleviate suffering is accompanied by thoughtful and appropriate action, which goes beyond just sharing the experience of another person’s emotional state.

Compassionate care is a good in itself - it is hard to imagine someone willingly choosing uncompassionate care for themselves or a loved one. It is also beneficial for health outcomes. A literature review by Sinclair *et al* [2] reports that clinicians found compassion to be an effective medium for eliciting patient health information, and improving patient compliance and disclosure, while a hostile attitude towards patients was felt to diminish diagnostic accuracy and impinge medical decision-making. Patient-reported outcomes included increased sense of responsibility for and control over their health, increased hope, feeling heard and understood, and feeling more trust towards clinicians. The authors also noted the benefits of compassionate care for the clinicians themselves, such as increased job satisfaction and sustainment.

Leadership plays a key role in fostering compassionate healthcare. Indeed, the National Advisory Group on the Safety of Patients in England [3] explicitly places compassion among key elements of leadership:

> *’Culture change and continual improvement come from what leaders do, through their commitment, encouragement, compassion and modelling of appropriate behaviours’*. [3]

For an organisation to nurture a culture of compassion, its leaders must embody compassion in their leadership [4,5]. Moreover, as de Zulueta [6] has underlined, experiencing compassion makes people more able to show compassion to others, conducing to a virtuous spiral. Many models have been developed which seek to understand and define key leadership concepts. Two models have attracted particular attention in the healthcare sphere. The first is ‘servant leadership’ [7], because its focus upon the needs of others (altruism) and the setting, aside of egoistic goals, is arguably highly appropriate to the health care setting [6,8]. The other model is ‘resonant leadership’ [9], which sees compassion as an essential part of the concept of emotional intelligence upon which it is based.

A key requirement of compassionate care is that it must be culturally competent [1], this is fundamental in addressing the health inequalities which exist in culturally diverse societies, and leadership plays a key role in supporting effective training and implementation of this competence [10]. Nursing and other healthcare professions therefore require leadership that is both culturally competent and compassionate, which Papadopoulos [1] terms ‘culturally competent and virtuous leadership’ - an ethical activity founded upon compassion, honesty, kindness, fairness, altruism, and co-operation.

Fundamental to culturally competent and virtuous leadership is an appreciation of cross-cultural perspectives on both compassion and leadership. These are both culturally mediated [11,12], and cultures therefore differ in how these are conceived and expressed. However, there are also commonalities across cultures. For example, Robie *et al* [13] compared leadership in the USA and seven European countries, and suggested that there were more similarities than differences between them, while in their study of servant leadership Winston & Ryan [14] report that similar cultural concepts can be found in African, East Asian, Mediterranean, and Indian value systems. Cross-cultural perspectives on leadership are a key aspect of leadership research, with perhaps the most influential research programme on this topic being the GLOBE study [11], which examined leadership and organizational behaviour in 62 countries, and describes numerous instances of the relationship between cultural features and leadership styles. A cross-cultural archetype of leadership described in this study is humane-oriented leadership, which Winston & Ryan [14] suggest is a close parallel to servant leadership, and which, in turn, de Zulueta [6] suggests is closely aligned with the concept of compassionate healthcare leadership.

Akerjordet & Severinsson [15] note a lack of research concerning emotional intelligence in healthcare, and, until comparatively recently, there has been a corresponding lack of research into cross-cultural aspects of this topic. However, more recent research has made a start in understanding what aspects of compassion in healthcare management might apply across cultures. Kouta *et al* [16] report an attempt to develop a European model for healthcare leadership which promotes the values of cultural competence and compassion. Ten characteristics are needed by senior healthcare staff in order to provide compassionate and culturally competent leadership. In order of importance, these were defined as *Communicative, Fair, Compassionate, Encouraging, Organised, Friendly, Calm, Courageous, Independent*, and *Autonomous*. However, little is known about how these might be expressed behaviourally. This issue is important because, as discussed above, changes occur because of what leaders do - the behaviours, attitudes, and personal qualities that they model. Furthermore, little is known, about how the expression of culturally competent and virtuous healthcare leadership might differ between different cultures, or about whether there might be core behavioural attributes (i.e. key indicators of compassionate leadership) which apply across cultures. In an attempt to address this gap in our knowledge, this paper explores what behaviours an international group of nursing and midwifery managers thought characterised compassionate leadership. The findings are discussed in relation to the GLOBE study’s humane-oriented leadership archetype, and Kouta *et al’s* [16] findings concerning the elements of culturally competent and compassionate healthcare leadership.

## METHODS

### Design

The study was a cross-sectional descriptive survey of the views and experiences of nursing and midwifery managers concerning compassion in their work setting.

### Procedures

The survey was developed in partnership with collaborators in the participating countries and included closed and opened-ended questions. This paper focusses solely upon the responses to the open-ended survey question concerning which actions and behaviours indicated that a manager was exercising compassionate leadership.

The survey questions were translated, pre-tested, and checked for clarity by the international team. The translation and back-translation into English of the survey, and the translation of the completed surveys into English, were conducted following the World Health Organisation (WHO) guidelines [17]. All locally-collected data were translated into English by the international collaborators before the completed surveys were compiled and sent for analysis.

The two inclusion criteria for participation were that participants should be a registered nurse or midwife, and hold a managerial position in a hospital, community, educational, or other setting. A snowball sampling method was used, with a minimum of 40 participants to be recruited from each country for it to be included. Data collection took place between the end of November 2017 and the end of July 2018, with the data being collected using the web-based electronic survey software *Qualtrics*. Potential participants were sent the link to the survey along with an invitation letter explaining the aim and procedures of the study. Participation was anonymous, confidential, and completely voluntary, and completion of the survey constituted consent to the study.

### Sample

A convenience sample of 1,217 participants was recruited from 17 countries. The sample was predominantly female (85%), 67% of the participants had more than five years’ management experience, and 45% managed more than 20 staff. Most participants (64%) reported working in a hospital setting, 14% in an educational setting, 13% in community settings or primary care, and 10% worked in other settings.

The main open-ended question received a total of 933 responses, with a mean of 60 responses per country (range = 33 to 110 responses). From the 1,217, 284 participants did not answer this particular question and therefore could not be included in the analysis. The missing data were found to be evenly distributed among the countries and no statistically significant relationship was found between the number of responses and the nurse/midwife density for each participating country. The number of responses and the most recent WHO data concerning nurse/midwife density (number of nurses/midwives per 10,000 population) [18] for each country were entered into an *IBM SPSS* (version 25) database, and a Pearson’s correlation found a non-significant correlation coefficient of r = -0.187, p=0.472 between these variables.

### Analysis

Content analysis was used to analyse the responses. Firstly, one author read each response and extracted significant statement(s) from it, as described in [19]. Each extracted statement was then assigned to a category which was given a descriptive label reflecting its meaning, with new categories being created as the work progressed.

Because more than one statement could be extracted from each response, 1,609 statements were assigned to 54 categories by the end of the process, with a mean of 30 statements per category (range = one statement to 212). A number of categories were made up of comparatively few responses, so in order to achieve a smaller, more manageable number of categories the 19 categories in the first quartile of the distribution (which each had fewer than 14 responses) were removed. The 35 remaining categories accounted for 1,298 responses, or 93% of the original total. A further examination of the coding after the analysis was complete found that data saturation had been reached after 802 responses (89%) had been analysed.

The 35 categories were then treated as sub-themes, and three of the authors consequently collaborated in inductively sorting them into groups based upon their similarity to a common overarching idea, or theme, which were each given descriptive labels also. The themes which had emerged from this process (in italic), and the sub-themes (capitalised, and in inverted commas) which constitute them, are presented in Table 1.

**Table 1.** Themes and sub-themes (quartiles 2 to 4)

| **Theme (% statements)** | **Sub-themes (n statements)** |
| --- | --- |
| *Virtuous Support* (36%) | 1. Aware of and Flexible Towards Individual Staff Members' Circumstances (emergencies, etc.) (178)<br>2. Actively Supports Staff When Necessary/Acts as Staff Advocate (152)<br>3. Acts to Solve Problems/Resolve Conflicts (130)<br>4. Takes an Interest In and Knows the Members of the Staff Team (59)<br>5. Amenable/Usually Responds Favourably to Reasonable, Non-urgent Requests (21) |
| *Communication* (27%) | 1. Sensitive, Active Listener (212)<br>2. Available/Approachable/Open/Makes Time to Listen (120)<br>3. Actively Promotes Communication (49)<br>4. Staff Team Can Communicate Openly With the Manager (24) |
| *Personal Virtues of the Manager* (17%) | 1. Empathic/Understanding/Accepting/Non-judgemental/Warm (151)<br>2. Considerate/Attentive/Kind/Amiable (70)<br>3. Is Fair (29) |
| *Participatory Collaboration* (7%) | 1. Collaborative/Open to Feedback and Ideas/Compromises/Negotiates (64)<br>2. Works Alongside Staff/Leads Rather Than Dictates (21)<br>3. Takes Questions/Suggestions/Concerns of Staff Seriously (18) |
| *Growth/Flourishing/Nurturing* (7%) | 1. Gives Positive Feedback and Praise (27)<br>2. Appreciative/Makes the Team Feel Valued (23)<br>3. Sensitive, Tactful Handling of Mistakes/Avoids Blame Culture (21)<br>4. Encouraging/Motivates and Empowers the Team (16)<br>5. Supportive of CPD/Training/Growth (15) |
| *Team Cohesion* (6%) | 1. Mutual Respect Between Manager and Staff Team (45)<br>2. The Team is Happy Within Itself/Good Relationships Within the Team (32)<br>3. Active Focus Upon and Promotion of Teamwork (17) |

After the completion of the coding and the identification of sub-themes and themes, the percentage of responses contained within each theme were calculated, this indicated the contribution made by each theme’s responses to the overall total. Displayed as a stacked bar chart, this produced a visual composite of the views expressed in the overall responses. Similar charts were produced for each country, which enabled them to be compared to the profile of the overall results, and to each other. These profiles are presented in Figure 1.

**Figure 1.**
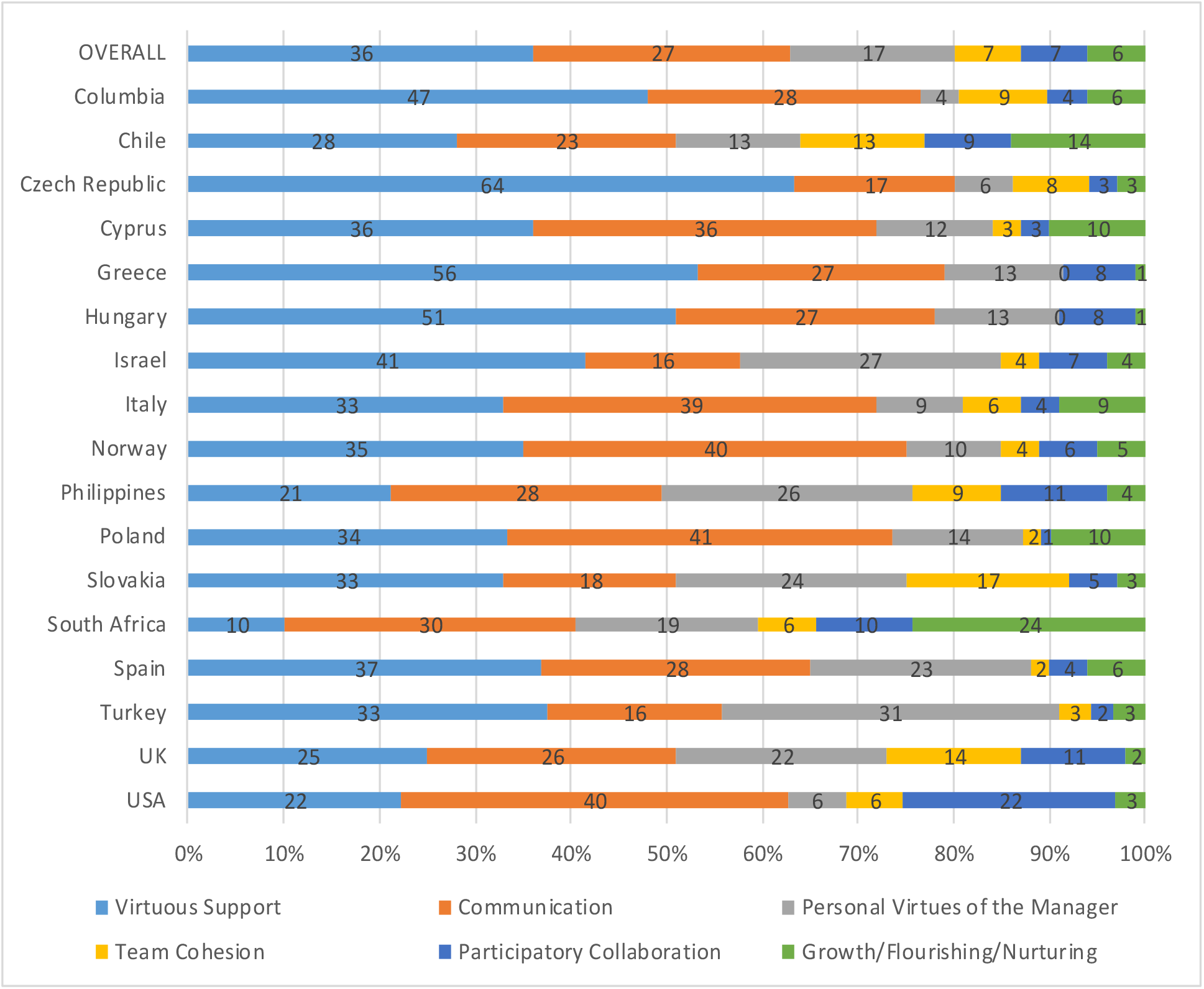
Overall and national profiles of compassionate nursing/midwifery management themes (quartiles 2-4)

The same process was repeated on the sub-themes in the fourth quartile of the response distribution alone, which had each received between 68 and 212 responses. The fourth quartile contained seven sub-themes, and the number of themes was reduced from six to three. These seven sub-themes accounted for 1,013 responses. Since 1,609 statements had originally been derived from the participants’ responses and classified into 54 sub-themes, this meant that 13% of the sub-themes accounted for 63% of the statements that had been extracted from the participants’ responses, and can therefore be regarded as representing what the participants regarded as the most important characteristics of compassionate management behaviour. These themes and sub-themes are presented in Table 2, and the profiles derived from them are presented in Figure 2.

**Table 2.**
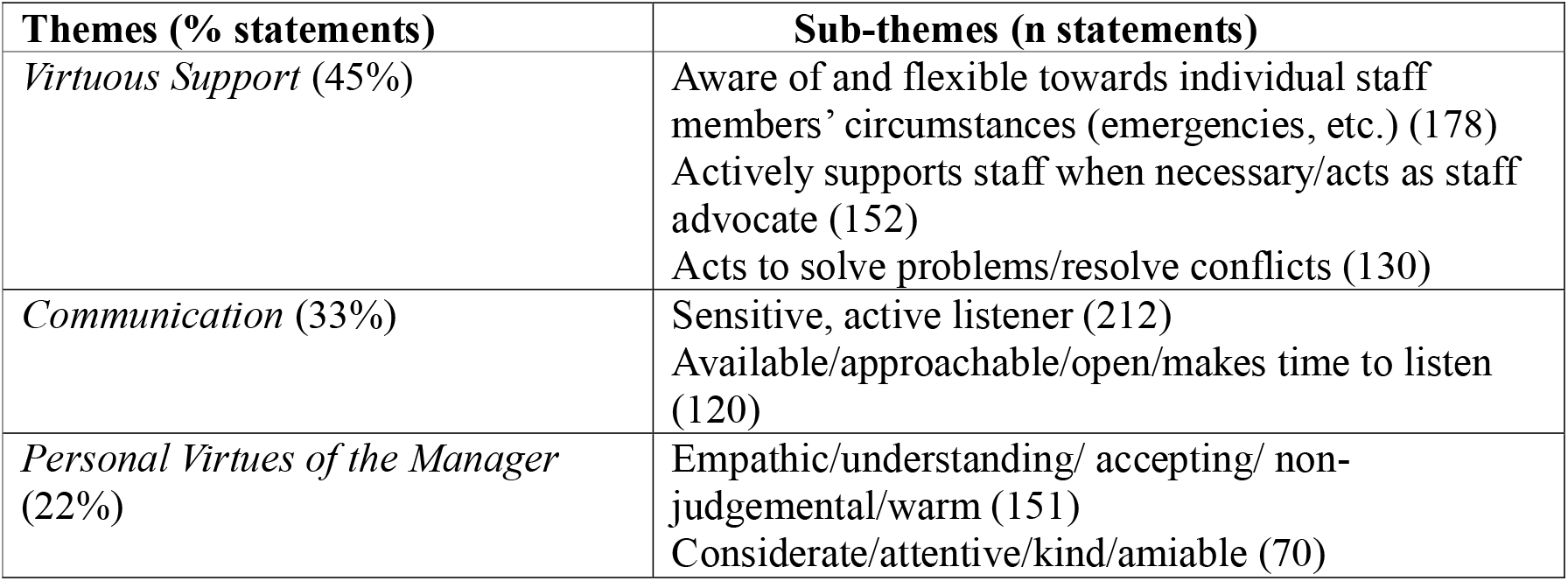
Themes and sub-themes (quartile 4 only)

**Figure 2.**
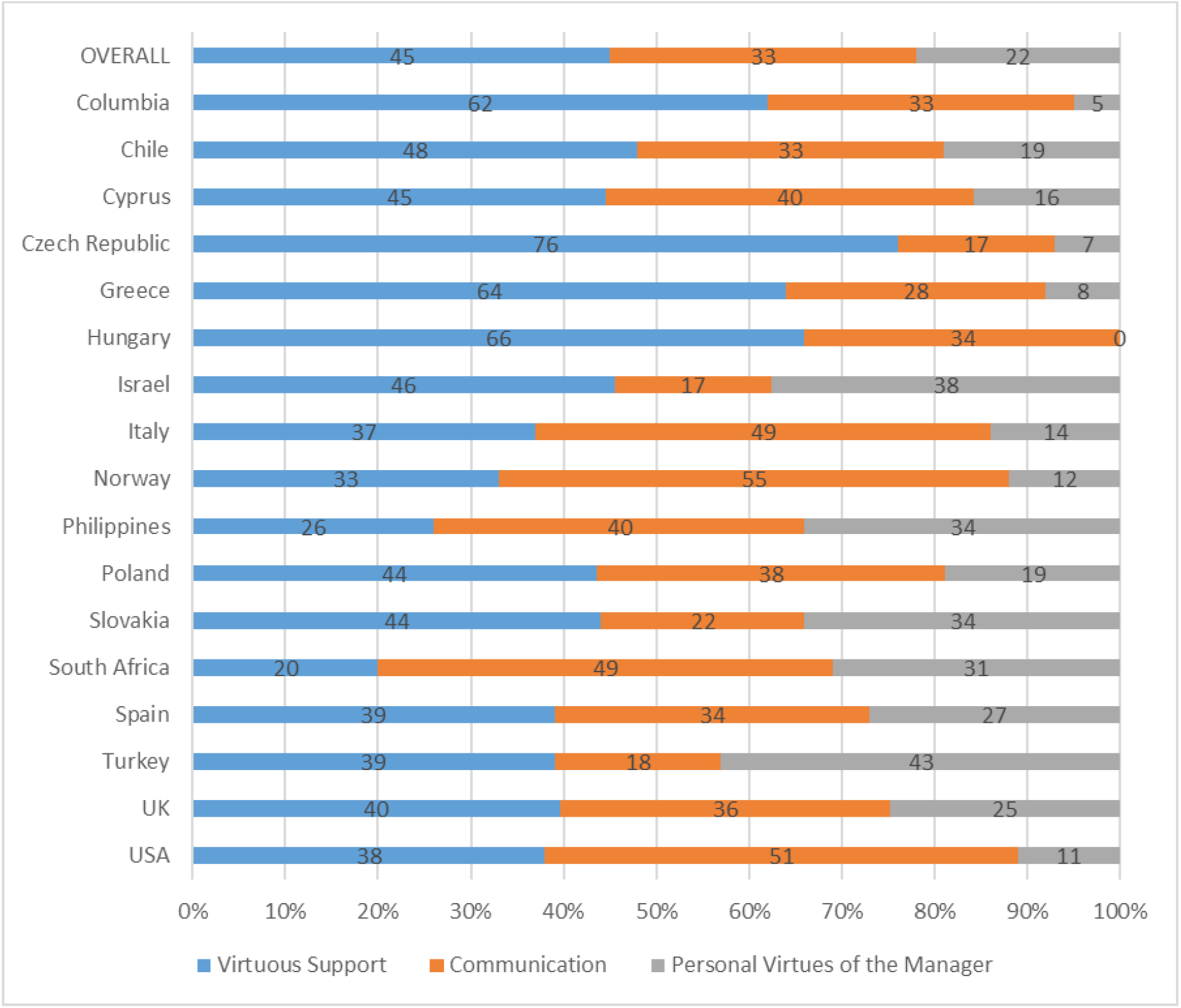
Overall and national profiles of compassionate nursing/midwifery management themes (quartile 4 only)

## RESULTS

In the overall profile the theme *Virtuous Support* accounted for most of the responses. The Czech respondents scored the highest on this group, followed by Hungary, Greece, and Columbia, whilst South Africa scored the lowest, followed by the Philippines. The sub-themes contained within *Virtuous Support* include being sympathetic to and supportive of subordinates with health- or childcare-related problems, acting supportively of their team (which may include advocacy), and problem solving (including conflict resolution). The term *Virtuous* is used to reflect the Aristotelian definition of moral virtue as a disposition to behave in the right manner, which strikes a balance between the extremes of deficiency and excess, which are seen as vices. *Virtuous Support* then, refers to the compassionate manager’s steady and consistent disposition to do the right thing by his/her staff.

The responses in the next-largest theme, *Communication*, portray a manager who actively promotes open communication between themselves and their team, who is available and approachable, who makes time to listen to his/her staff members, and listens well. Respondents from Poland scored highest on this group, followed by Norway, the USA, Italy and Cyprus, with respondents from Slovakia scoring lowest, followed by Israel and Turkey. It is noteworthy that while *Communication* is the theme with the second-highest number of responses, it contains the sub-theme which accounts for the highest number of responses (’Sensitive, Active Listener’), which suggests the importance ascribed to it by the participants for culturally competent and virtuous leadership.

*Personal Virtues of the Manager* represents a disposition towards empathy, warmth, kind consideration, and fairness in the compassionate manager, and is largely analogous to the concept of unconditional positive regard [20]. The highest-scoring country for *Personal Virtues of the Manager* was Turkey, followed by Israel and the Philippines, while the lowest score was from Columbia, followed by the USA and Czech Republic.

For the remaining themes, the USA scored highest on *Participatory Collaboration*, followed by the UK and the Philippines, with South Africa scoring highest on *Growth/Flourishing/ Nurturing*, followed by Chile, with Greece and Hungary scoring lowest, and Turkey scored highest on *Team Cohesion*, with Cyprus and the Czech Republic scoring lowest. However, these last three themes only accounted for 20% of the responses, and none of their sub-themes were represented in the fourth quartile of the responses.

## DISCUSSION

Our findings indicate that the themes of Virtuous Support, Communication, and Personal Virtues of the Manager can be seen as the most important elements of culturally competent and virtuous leadership reported by the participants. As such it may be of use in informing planning for training in it, and offer at least a starting point in understanding cross-cultural differences and similarities in compassionate leadership behaviour. These key elements can be summarised as follows:

1. Be someone who others feel comfortable speaking to, and listen well when they do.
2. Be sympathetic and helpful if unexpected events happen to members of your staff (especially those with child- or other care responsibilities).
3. Actively support your staff team, speaking up for them when necessary.
4. Actively help to resolve problems and conflicts.

Being empathetic with your staff was also reported by nursing managers in Japan as an important attribute in their efforts to achieve change in their wards [21].The themes and sub-themes relating to the behavioural expression of compassionate healthcare management represented in our findings are consistent with findings concerning the qualities of compassionate healthcare management reported in Kouta *et al* [16]. The qualities *Communicative, Fair* and *Friendly* correspond closely with our themes and sub-themes of *Communication*, ‘Is Fair’ and ‘Considerate/Attentive/ Kind/Amiable’ (from *Personal Virtues of the Manager)* respectively. Kouta *et al*’s *Compassion* and *Courageous* have no direct correspondence within our findings, but they are implicit within the sub-themes which comprise *Virtuous Support*. Indeed, the word *Virtuous* was included in this theme’s label precisely in order to represent the courage that is often required in supporting and advocating for the staff team. Again, Kouta *et al*’s *Calm* is not replicated in our findings as such, but is implicit to a certain extent in the sub-theme ‘Sensitive, Tactful Handling of Mistakes/Avoids Blame Culture’ (from *Growth/Flourishing/ Nurturing*).

In relation to the GLOBE, the categories that make up Virtuous Support and Personal Virtues of the Manager echo the GLOBE’s definition of humane leadership orientation which is comprised of supportiveness, compassion, generosity, and modesty [11]. However, there are inconsistencies with the clustering of countries and their scoring. Countries participating in the GLOBE study are organised into ten clusters based upon purported cultural similarities. The clusters which score highly on humane orientation are listed as South Asia, Sub-Saharan Africa, and Anglo, while the lowest scoring are the Latin Europe and Nordic Europe clusters [11]. The highest scoring countries on *Virtuous Support* in our sample were (in descending order) the Czech Republic, Greece, Hungary, and Columbia. Columbia is part of the Latin America cluster, whereas Greece and Hungary are in the Eastern Europe cluster, and while the Czech Republic is not included in the GLOBE study, it shares borders with Slovakia and Poland which are in that cluster; however, neither the East Europe nor the Latin America cluster score highly on humane orientation.

Conversely, South Africa (Sub-Saharan Africa), the Philippines (South Asia), the USA (Anglo), and the UK (Anglo) scored lowest on *Virtuous Support*, while their respective country clusters are listed in GLOBE as scoring highly on humane orientation. A similar inconsistency was seen in the results for *Personal Virtues of the Manager*, where none of the countries scoring highly (Turkey, Israel, Slovakia) belong to GLOBE’s high humane orientation country clusters, and none of the low-scoring countries for *Personal Virtues of the Manager* (the Czech Republic, the USA, and Columbia) belong to low humane orientation country clusters.

Whilst the GLOBE study is often referred to as the benchmark for the cultural /country comparisons for the differences in humane leadership, we found that their findings did not provide useful insights for the interpretation of ours. This is largely due to the fact that their sample populations were very different from ours as they used managers of food, financial, and telecommunications industries whilst we used healthcare managers, primarily in nursing and midwifery. It is important to continue the exploration of cross-cultural understanding of nursing leadership behaviours since other researchers continue to find differences among cultures. For example, Boldy *et al* [22] reported that Western Australians found a transformational leader as an effective nurse manager, whereas responders from Tanzania found a transactional leader as an effective nurse manager, and those from Singapore referred to a leader with a combination of attributes of both leadership styles. The authors concluded that healthcare leadership development courses need to include the cultural dimension in order to be effective.

Our study made no attempt to determine adequate sample size. However, this was not an important consideration because the aim of the study was to explore managers’ views rather than to test hypotheses, so the issue of statistical power was not relevant. Vasileiou *et al* [23] discuss the issue of sample size sufficiency in interview-based research (which is comparable to our study, given the nature of our data). They found that the median number of interviews in qualitative papers published in three medical journals, over 15 years, were 15, 30.5, and 31 respectively, which suggests that our inclusion requirement of at least 40 responses per country gave an ample sample size. This conclusion is further supported by our finding that data saturation had been achieved after 89% of the responses had been analysed.

The selection of the countries involved was influenced by the networks of the first author. This accounts for the Eurocentric bias of the sample, with few countries from Africa, Asia and the American continent represented within it. The findings are also vulnerable to the usual biases that are associated with self-report, highlighted in Taras *et al* above [24]. A final source of potential bias is the fact that the number of responses from each country was unrelated to the density of nurses and midwives in their respective populations. While the density of nurses and midwives in a country’s population might not correspond exactly with the density of nursing and midwifery managers, we cannot rule out the possibility that the true range of opinions and views is under-represented among countries with relatively fewer participants, or that the views from countries with relatively more participants exert disproportionate influence. This emphasises the need for international surveys of this kind to stratify each participating country’s sample in relation to their respective nursing/midwifery density.

This study adopted a country-based definition of culture. While this method is unsatisfactory in some ways, as it does not address the diversity of cultures within countries and thus the complexity of the study participants’ cultural identities, it has been used in other cross-cultural studies, notably by Hofstede [25] and the GLOBE study itself [11]. This issue is worthy of further research and should bear consideration in planning cross-cultural research.

## CONCLUSIONS

Our findings identified key aspects of compassionate management in nursing and midwifery which might be useful in planning training in healthcare management. The method used to profile our results went beyond presenting a narrative account of the themes that emerged from the survey responses, and provided a striking and direct way of comparing the responses of each country with both each other and with the norm for the whole sample. While at first sight it may seem unusual to represent qualitative data in numerical form, other qualitative approaches, for example Q-sort, Delphi Method, and repertory grids, all apply quantitative analysis to qualitative data in order to systematically examine different perspectives on the issue in question. It is axiomatic that culturally competent and virtuous leadership in nursing and midwifery must be informed by cross-cultural perspectives on both compassion and leadership. However, our findings emphasise that these cannot be adopted uncritically from other disciplines.

## Data Availability

Data availability upon request to the corresponding author

## Authorship contributions

Study design: IP

Data collection: ALL AUTHORS

Data analysis: IP, SW, RL, CK

Study supervision: IP

Manuscript writing: IP, SW, RL, CK

Critical revisions for important intellectual content: IP

## Acknowledgments

We thank all the 1,217 participants who gave their time to complete the survey. We would like to additionally thank Dr Tommaso Batistoni, Sheila Ali, Syed Miah, and Dr María José Morales Gázquez for their contributions to the data analysis process.

## Key Messages

- Main key attributes of a compassionate leader were found from a large international sample of nurses and midwives.
- These key attributes are: *Virtuous Support, Communication, Personal Virtues of the Manager, Participatory Communication, Growth/Flourishing/ Nurturing*, and *Team Cohesion*.
- Despite the cross-cultural variations, these key attributes were widely represented among different countries’ responses and may be useful in informing changes in policy, training and practice.

